# Large language models for abstract screening in systematic- and scoping reviews: A diagnostic test accuracy study

**DOI:** 10.1101/2024.10.01.24314702

**Authors:** Christian Hedeager Krag, Trine Balschmidt, Frederik Bruun, Mathias Brejnebøl, Jack Junchi Xu, Mikael Boesen, Michael Brun Andersen, Felix Christoph Müller

**Affiliations:** Copenhagen University Hospital, Herlev and Gentofte; Radiological AI Testcenter (RAIT.dk); Copenhagen University Hospital, Bispebjerg and Frederiksberg Hospital

## Abstract

**Introduction:** We investigated if large language models (LLMs) can be used for abstract screening in systematic- and scoping reviews.

**Methods:** Two broad reviews were designed: a systematic review structured according to the PRISMA guideline with abstract inclusion based on PICO criteria; and a scoping review, where we defined abstract characteristics and features of interest to look for. For both reviews 500 abstracts were sampled. Two readers independently screened abstracts with disagreements handled with arbitrations or consensus, which served as the reference standard. The abstracts were analysed by six LLMs (GPT-4o, GPT-4T, GPT-3.5, Claude3-Opus, Claude3-Sonnet, and Claude3-Haiku). Primary outcomes were diagnostic test accuracy measures for abstract inclusion, abstract characterisation and feature of interest detection. Secondary outcome was the degree of automation using LLMs as a function of the error rate.

**Results:** In the systematic review 12 studies were marked as *include* by the human consensus. GPT-4o, GPT-4T, and Claude3-Opus achieved the highest accuracies (97% to 98%) comparable to the human readers (96% and 98%), although sensitivity was low (33% to 50%). In the scoping review 130 features of interest were present and the LLMs achieved sensitivities between 74-84%, comparable to the human readers (73% and 86%). The specificity of GPT-4o (98%) and GPT-4T (>99%) greatly surpassed the other LLMs (between 33% and 93%). For abstract characterization all LLMs achieved above 95% accuracy for language, manuscript type and study participant characterisation. For characterisation of disease-specific features only GPT-4T and GPT-4o showed very high accuracy. For abstract inclusion the highest automation rate (91%) at the lowest error rate (8%) was achieved by use of two LLMs with disagreement solved by human arbitration. An LLM pre screening before human abstract screening achieved an automation rate of 55% with no missed abstracts.

**Conclusion:** Abstract characterisation and specific feature of interest detection with LLMs is feasible and accurate with GPT-4o and GPT-4T. The majority of abstract screenings for systematic reviews can be automated with use of LLMs, at low error rates.

## Introduction

Systematic reviews and scoping reviews are methods used for evidence synthesis. The Cochrane networks have defined systematic reviews as reviews “*of a clearly formulated question that use systematic and explicit methods to identify, select, and critically appraise relevant research, and to collect and analyse data from the studies that are included in the review*”^1^. In comparison, scoping reviews are “*exploratory projects that systematically map the literature available on a topic, identifying key concepts, theories, sources of evidence and gaps in the research*”, according to the definition by the Canadian Institute of Health Research^2^.

Systematic and scoping reviews rely on a proper search strategy, title and abstract screening, full text assessment, and finally extraction of variables from the included papers. Human readers usually do this manually, often following guidelines, for example, the PRISMA guidelines for systematic reviews^3^.

Abstract screening is necessary because full text assessment of all records for larger systematic reviews would be impractical and unnecessary. But, because abstracts excluded during screening do not undergo full text assessment, the step is critical. One of the issues with manual abstract screening and extraction is varying inter-reader reliability. Inter-reader reliability between human readers in systematic reviews is rarely very good, and can range between κ = 0.37-0.90^4,5^. These inter-reader variabilities are primarily caused by human errors, where relevant information is missed from the title or abstract^4^. Conducting repetitious work for several hundreds of hours can leave humans prone to errors. It has previously been described that individual coder performance is affected by “learning effect” and “fatigue effect”^6^. To reduce the number of abstracts which have to be screened, narrower search strategies are sometimes conducted to limit the amount of work. This strategy is not without pitfalls, as narrower search strategies may overlook relevant papers. Finally, the fast-growing number of publications challenges the systematic review process. A large-scale systematic review can take more than a year to conduct and cost up to a quarter of a million USD^7,8^. Currently PubMed alone contains more than 37 million citations of biomedical literature from MEDLINE, life science journals, and online books^9^.

How could we reduce the workload on reviewers for abstract screening, while maintaining the broadest possible search strategy? Large language models (LLMs) may be the answer to systematic and scoping review automation^10^. Researchers assisted by LLMs could conduct extensive reviews of hundreds of thousands to millions of studies - ensuring that each abstract is evaluated. In the literature, there are already beginning to appear examples of how LLMs can be used for systematic reviews^11–15^. We wanted to explore this subject further with multiple different LLMs from different companies, expanding the concept to both systematic and scoping reviews and estimate the workload reduction.

Our primary research question was to determine the diagnostic test accuracy of six commercially available LLMs for abstract characterisation, screening abstracts for inclusion and detection of specific features using a zero-shot prompting approach. Our secondary and tertiary research questions were to determine the number of abstracts that could be automatically screened using LLMs and to determine the intra- and inter-reader agreement of different LLMs. Our hypothesis was that LLMs could perform these tasks with accuracies and agreements close to that of humans.

## Methods

### Study design

The study design is a prospective, diagnostic test accuracy study of LLMs. We examined the diagnostic test accuracy of multiple LLMs extracting information from abstracts for two review studies (Table 1), a systematic review and a scoping review. Data collection for the two reviews was planned before the index test and reference standard were performed. We followed the STARD guidelines for writing the manuscript^16^.

**Table 1:**
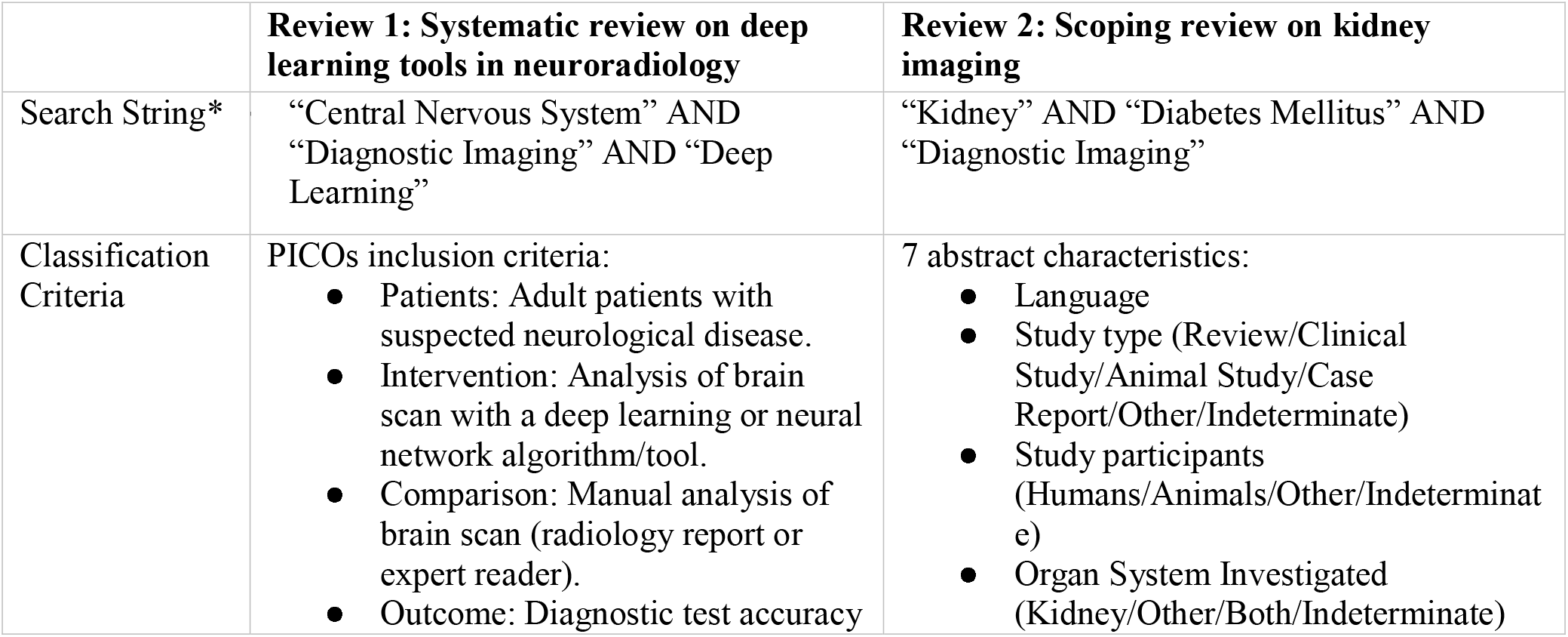
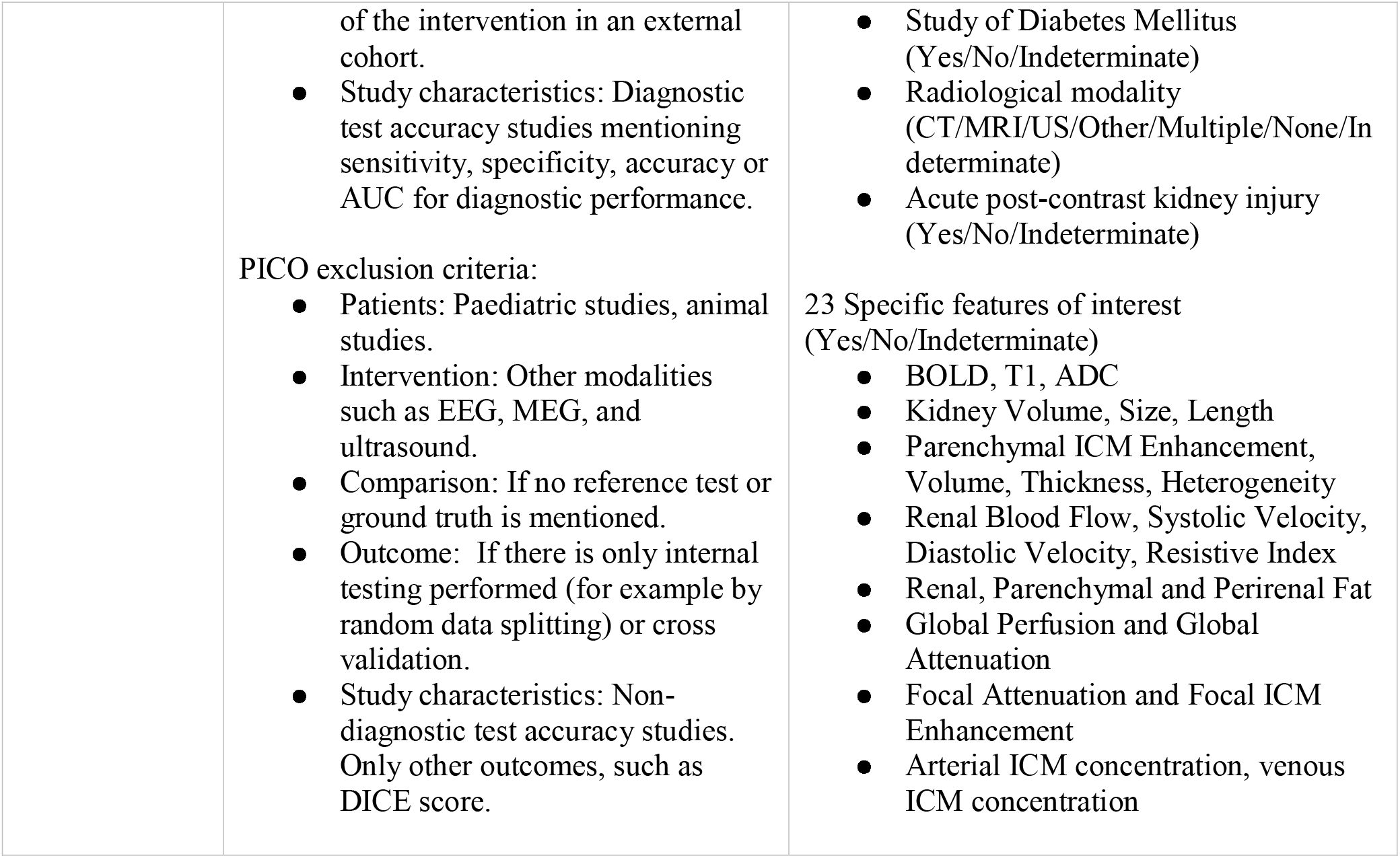
Classification criteria for the two reviews. ^*^Shortened, full search strings in supplementary materials.

|  | **Review 1: Systematic review on deep learning tools in neuroradiology** | **Review 2: Scoping review on kidney imaging** |
| --- | --- | --- |
| Search String* | “Central Nervous System” AND<br>“Diagnostic Imaging” AND “Deep Learning” | “Kidney” AND “Diabetes Mellitus” AND<br>“Diagnostic Imaging” |
| Classification Criteria | PICO's inclusion criteria:<br>● Patients: Adult patients with suspected neurological disease.<br>● Intervention: Analysis of brain scan with a deep learning or neural network algorithm/tool.<br>● Comparison: Manual analysis of brain scan (radiology report or expert reader).<br>● Outcome: Diagnostic test accuracy | 7 abstract characteristics:<br>● Language<br>● Study type (Review/Clinical Study/Animal Study/Case Report/Other/Indeterminate)<br>● Study participants (Humans/Animals/Other/Indeterminate)<br>● Organ System Investigated (Kidney/Other/Both/Indeterminate) |
|  | of the intervention in an external cohort.<br>Study characteristics: Diagnostic test accuracy studies mentioning sensitivity, specificity, accuracy or AUC for diagnostic performance.<br>PICO exclusion criteria:<br>Patients: Paediatric studies, animal studies.<br>Intervention: Other modalities such as EEG, MEG, and ultrasound.<br>Comparison: If no reference test or ground truth is mentioned.<br>Outcome: If there is only internal testing performed (for example by random data splitting) or cross validation.<br>Study characteristics: Non-diagnostic test accuracy studies. Only other outcomes, such as DICE score. | Study of Diabetes Mellitus (Yes/No/Indeterminate)<br>Radiological modality (CT/MRI/US/Other/Multiple/None/Indeterminate)<br>Acute post-contrast kidney injury (Yes/No/Indeterminate)<br>23 Specific features of interest (Yes/No/Indeterminate)<br>BOLD, T1, ADC<br>Kidney Volume, Size, Length<br>Parenchymal ICM Enhancement, Volume, Thickness, Heterogeneity<br>Renal Blood Flow, Systolic Velocity, Diastolic Velocity, Resistive Index<br>Renal, Parenchymal and Perirenal Fat<br>Global Perfusion and Global Attenuation<br>Focal Attenuation and Focal ICM Enhancement<br>Arterial ICM concentration, venous ICM concentration |

### Participants

For the systematic review (Review 1), we designed a review of the external performance of deep learning tools in neuroradiology following the PRISMA guidelines with a research question defined through PICOs criteria. For the scoping review (Review 2), we designed a review which would provide an overview of the literature, including abstract characteristics and imaging biomarkers used (specific features of interest), of diabetes-related kidney changes.

### PubMed search and extraction of abstracts

We conducted broad PubMed searches for relevant studies, and extracted date, DOI, authors, title, and abstract from PubMed with a Python extension (Biopython version 1.83). We conducted the systematic review search on April 19th, 2024 and the scoping review search on May 14th, 2024. For both reviews, we took out a sample of 500 abstracts for manual review (Figure 1).

**Figure 1:**
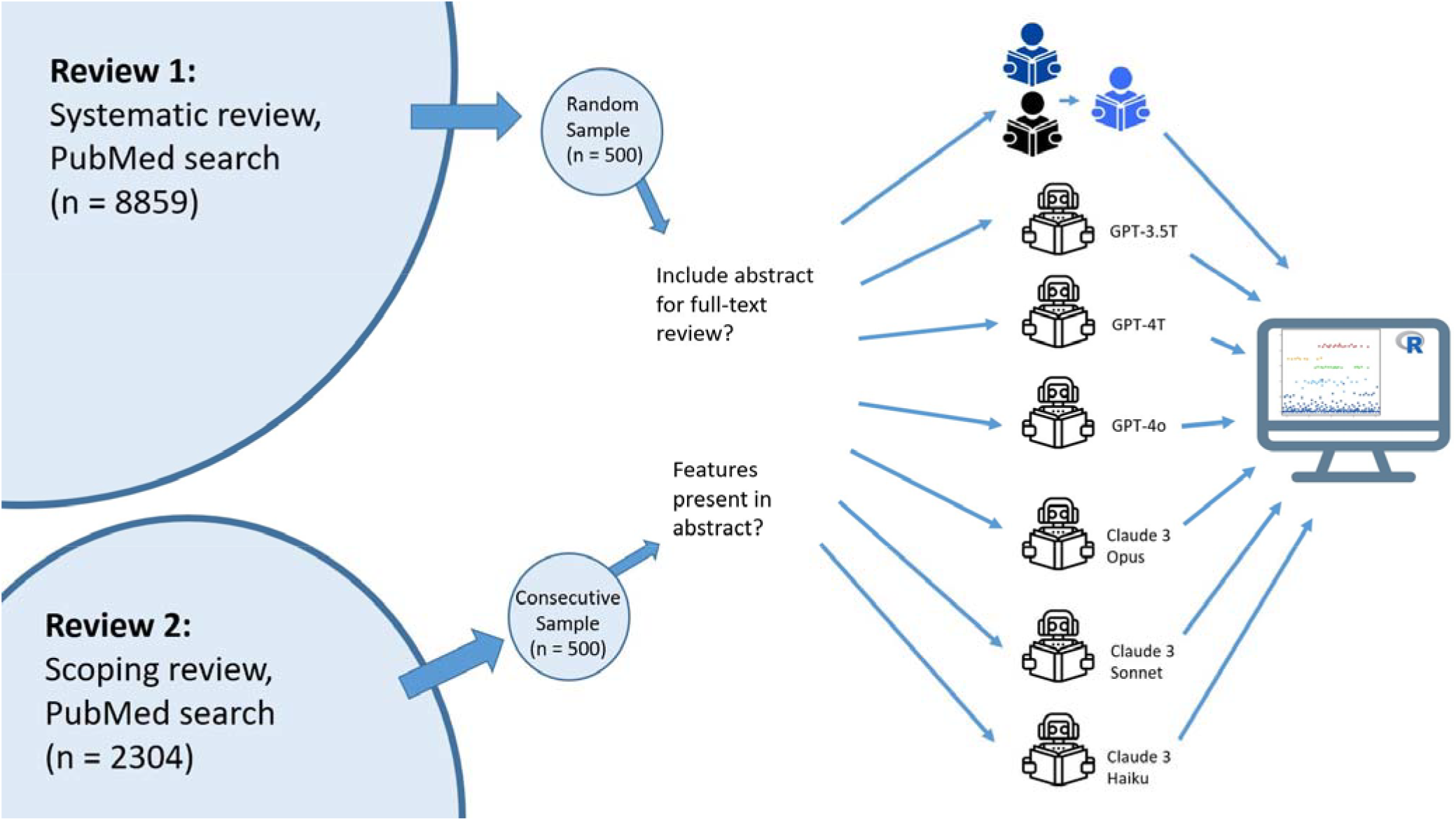
Study design

### Outcome measures

For the systematic review each abstract screened could be categorised as *included* in full-text review, *excluded*, or *indeterminate. Indeterminate* results would, according to PRISMA, also undergo full-text review and were therefore categorised as *included*.

For the scoping review, each abstract was assessed regarding 7 abstract characteristics (see Table 1). In addition, a list of 23 specific features of interest (see Table 1), was precompiled from a systematic review of MRI biomarkers^17^, and extended by further characteristics specific to CT. Each specific feature of interest could be coded as *yes* (present), *no* (absent), or *indeterminate*. An abstract could contain multiple specific features of interest.

### Index test

Abstracts were analysed with six different commercially available LLMs with the same prompt and setup. The LLMs were prompted to return the output in a structured JSON format along the prespecified items which were then parsed into a table format. The LLMs used were three models with assumed lower parameter count: GPT-3.5 (gpt-3.5-turbo-0125), Claude3-Sonnet (claude-3-sonnet-20240229), and Claude3-Haiku (claude-3-haiku-20240307) and three frontier models, assumed to have higher parameter count: GPT-4T (gpt-4-turbo-2024-04-09), GPT-4o (gpt-4o-2024-05-13) and Claude3-Opus (claude-3-opus-20240229). Figure 1 illustrates the study design.

### LLM prompt strategies

For review 1, we designed the primary LLM-prompt to follow the PICOs inclusion and exclusion criteria. We also tested two other LLM-prompt designs: one that presented the LLMs with only the PICOs inclusion criteria, and another where the LLMs were asked to generate a confidence score of 0 to 100 for how well the criteria were fulfilled. For review 2, the LLMs were prompted to mark predefined characteristics as present, absent, or indeterminate. The full LLM prompts are available in the supplementary materials.

### Index results

The coded output of each of the LLMs served as the index result. For Review 1 any other output than *Exclude* was considered as *Include*. For the abstract characterisation in Review 2 if the LLMs output any other organs these were classified as *Other* and if the LLMs output any other specific radiological modality these were either classified as *Other* or *Multiple*. For the specific feature of interest extraction, any results other than *Yes* or *No* were categorised as *No*, indicating that the LLM did not find the specific feature of interest.

### Reference standard

#### Review 1

Before LLM-analysis, two radiology residents (CHK, TB) coded abstract inclusion manually in the program Covidence, blinded to each other’s evaluation, with a third reader (FCM) deciding in case of conflicting readings. The final list of excluded and included abstracts was defined as the reference standard. We chose this method, as it is the golden standard in systematic reviews and in accordance with the PRISMA guidelines. Furthermore it allowed us to explore human inter-reader agreements.

#### Review 2

Two radiology residents (FJB, TB) coded abstracts independently and manually according to the 7 abstract characteristics and 23 specific features of interest before LLM-analysis. Any disagreements were resolved by consensus reading.

### Primary outcome

For Review 1 the primary outcome was the sensitivity, specificity, and accuracy of correctly including an abstract in a full text review. An abstract classified as *Included* by both index and reference was considered true positive.

For Review 2 the primary outcome for abstract characterisation was the accuracy of the LLMs in correctly classifying each characteristic. For the specific features of interest, the primary outcome was the sensitivity and specificity of the LLMs in correctly identifying the presence of a specific feature of interest. A specific feature of interest classified as *Yes* by both index and reference was considered a true positive. All 23 features of interest were aggregated for the calculation of sensitivity and specificity.

### Secondary outcome

We investigated the potential automation rate of LLMs for systematic reviews - which we defined as the reduction in the number of abstracts to be screened by human readers. Four different LLM-assisted scenarios were modelled and compared against the default scenarios of two human readers screening all abstracts with a third reader to arbitrate in case of disagreement. In the *Single LLM reader* scenario, an LLM would be deployed as a second reader alongside a human reader, with arbitration conducted by another human reader. The *LLM pre-screen* scenario used an LLM as a prescreening tool before two human readers conducted the abstract screening with arbitration by a third human reader. The *LLM pre-screen + Single LLM reader* scenario combined both an LLM to prescreen and a different LLM to act as a second reader during abstract screening. Arbitrations were conducted by a third reader. Finally, in the *Double LLM reader* scenario two different LLMs conducted abstract screening with a human third reader arbitrating. The different scenarios are visualised in Figure 2. We reported the percentage of abstract screening reductions for human readers (automation rate), the number of incorrectly excluded abstracts (error rate) using data from Review 1.

**Figure 2:**
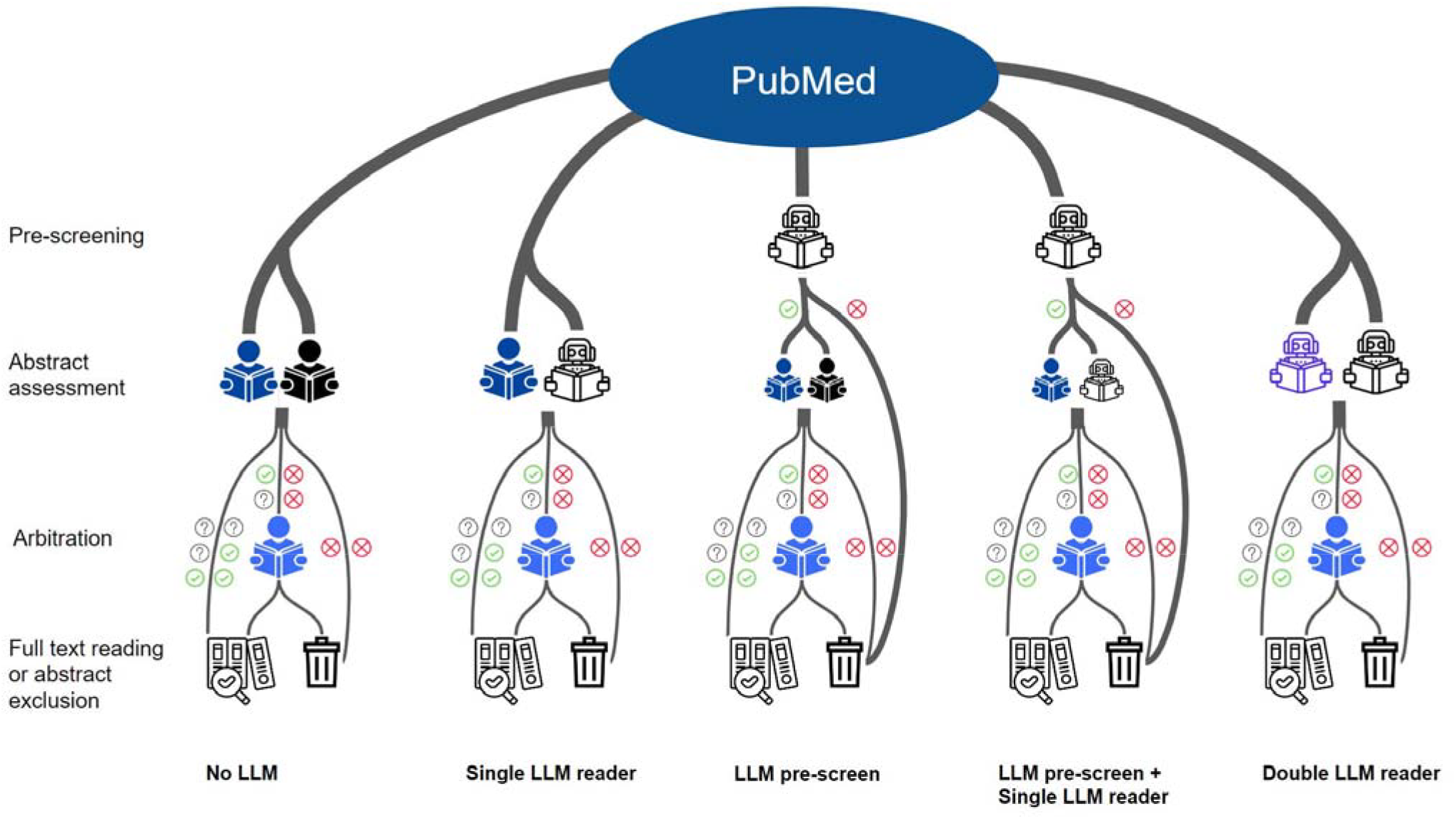
Proposal of LLM-assisted abstract screenings. From the left the standard human screening is pictured, followed by four scenarios of LLM-assistance in the abstract screening process.

### Tertiary outcomes

For both Review 1 and Review 2 we calculated inter-reader agreements between both human readers and LLMs and intra-reader agreements for the LLMs. Both analyses were conducted within a few weeks. For review 1, we also examined the effect of model temperature on the accuracy of the LLMs in review 1, two different prompt strategies and performed an in-depth qualitative analysis of all included abstracts.

### Statistical analysis

We conducted statistical analysis in R version 4.2.0 with the packages “tidyverse”, “caret”, “lme4”, “binom”, and “irr”. Sensitivity, Specificity, and Accuracy were calculated with 95% confidence interval using the Clopper and Pearson Binomial Exact Interval. Intra-reader and inter-reader agreements were calculated using Cohen’s Kappa. We analysed the differences between models and temperature settings across multiple runs with LLM-temperature setting at 0.2 and 0.8 using a generalised linear mixed model. The outcome variable was binary, defined as accurate diagnosis (yes/no), with the explanatory variables being the LLM model and LLM temperature.

## Results

### Review 1

The initial PubMed search returned 8859 unique abstracts, of which we took a random sample of 500 abstracts (5.6 %). Four abstracts and four DOIs were missing from the sample. Of the remaining 492 abstracts 12 were included for full-text review by the expert reader consensus. Input tokens averaged 768 per abstract (of which 325 comprised the prompt), output tokens averaged 107.6 per abstract per LLM. In total 17712 decisions on abstract inclusion were analysed with three different prompting strategies (prompt 1: 2952, prompt 2: 11808, prompt 3: 2952).

### Review 2

The initial PubMed search resulted in 2304 unique abstracts, and from them we took a consecutive sample of 500 (21.7%). Four abstracts were missing from the sample, and two abstracts did not have results from all LLMs resulting in a total of 494 included abstracts in the final cohort. Mean input tokens were 1218.5 for the content, prompt, and abstract and 294.6 for the output. Since LLMs were prompted twice, this resulted in 988 abstract summaries for each LLM containing 7 abstract characteristics (6916 per LLM; 41496 in total) and 23 specific feature of interest categorisations (22724 per LLM; 136344 in total). Incorrectly formatted output was returned for 28 of 494 (2.8%) abstracts by GPT-3.5, 7 of 494 (0.7%) abstracts by Claude3-Sonnet and 3 of 494 (0.3%) abstracts by GPT-4T, Claude3-Opus and Claude3-Haiku. GPT-4o returned all outputs in the correct format.

### Primary Outcome

#### Review 1

The three higher parameter models (GPT-4o, GPT-4T, Claude3-Opus) all showed specificities and accuracies above 97%, but low sensitivity for correct abstract inclusion in a systematic review. Their accuracy was on level with the accuracy of reader 1 and 2. The other three LLMs had moderate to high sensitivity but with low to moderate accuracy (table 2).

**Table 2:**
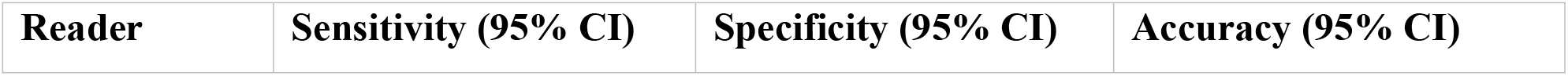
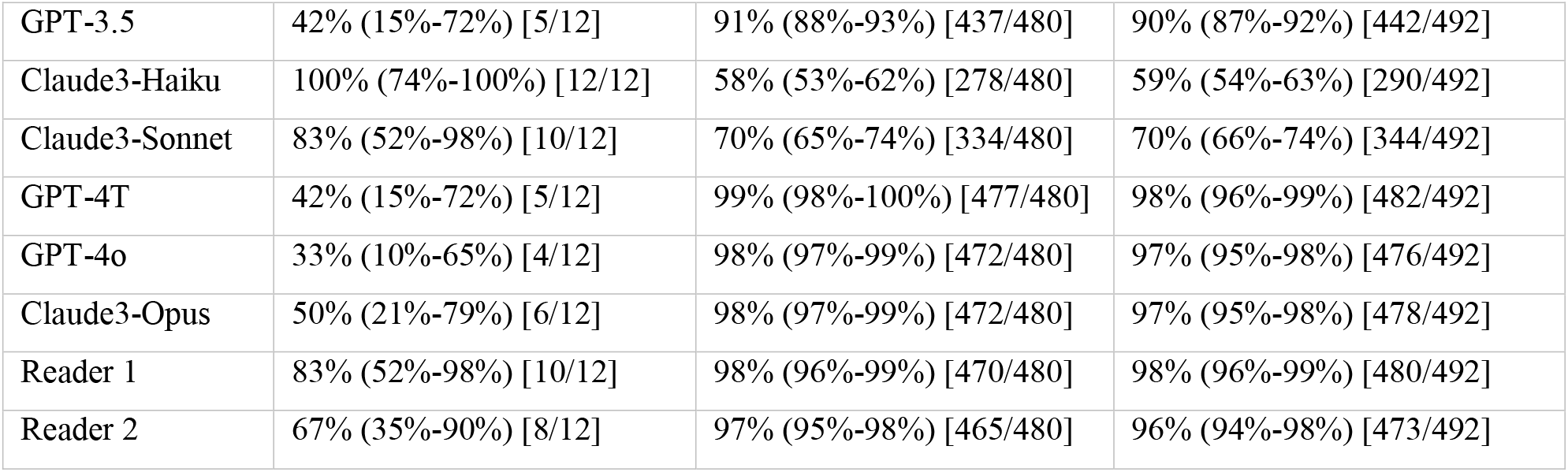
Diagnostic test accuracy of six LLMs for correct abstract inclusion for systematic review (review 1)

#### Review 2: Abstract Characterisation

All algorithms achieved accuracy above 95% for the classification of language, study type, and characterisation of human/animal study. For diabetes classification all higher parameter count LLMs achieved very high accuracy. For post contrast-acute kidney injury (PC-AKI) classification only GPT-4T and GPT-4o showed very high accuracy. For the classification of organ and modality, all LLMs demonstrated only moderate accuracy. Interestingly reader 2 also had low accuracy in the classification of the modality and reader 1 for the classification of the organ (Figure 3).

**Figure 3:**
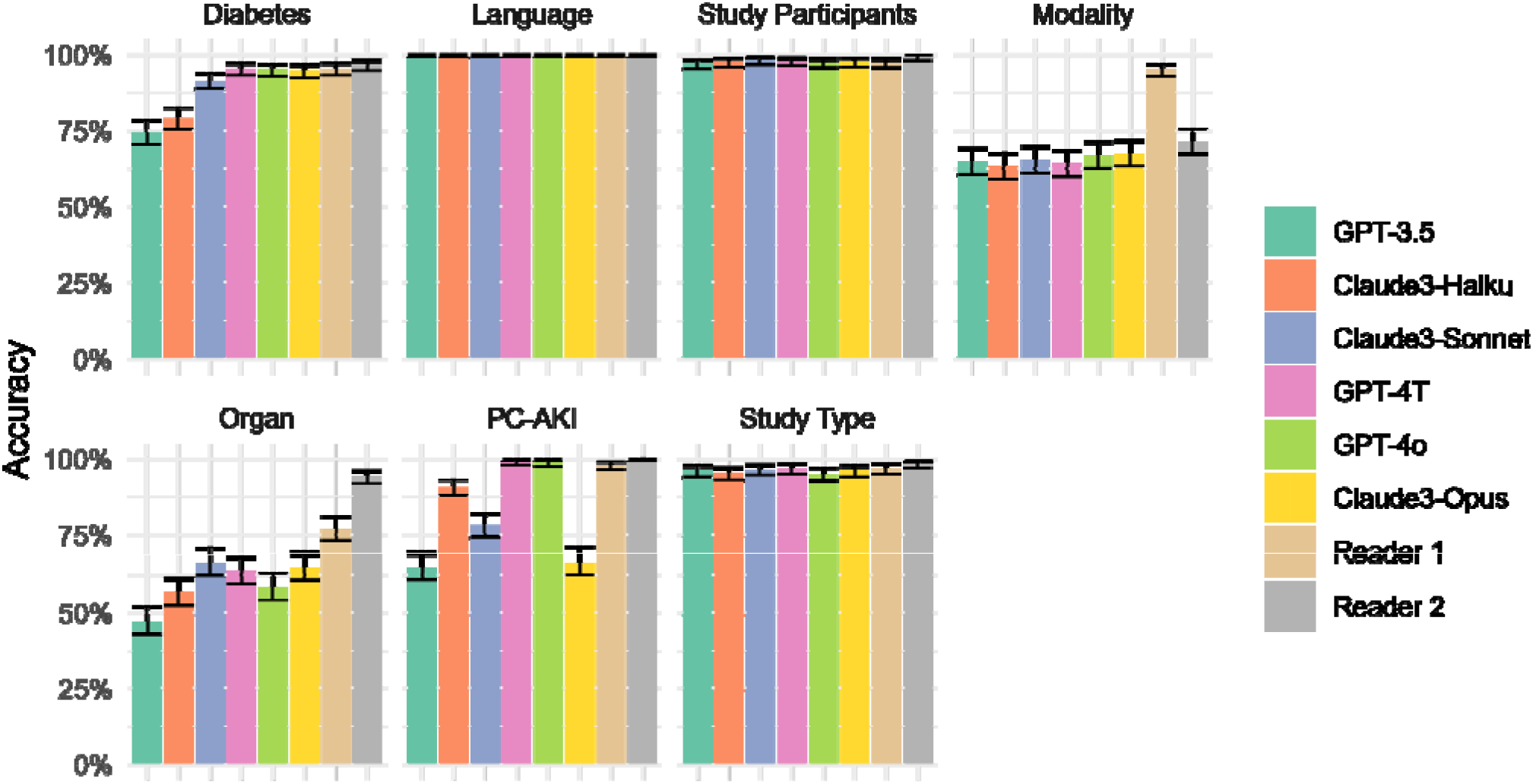
Accuracy of Large Language Models for Abstract Characterisation in Review 2, error bars show 95% confidence intervals using the binomial exact method. Human reader performance is shown for context.

#### Review 2: Special Feature of Interest Detection

For specific features of interest detection, all algorithms achieved a moderate to high sensitivity of between 74% and 84%, right between the sensitivity of the two reference readers (73% and 86%). GPT-4o and GPT-4T also had a very high specificity 98% and >99%, unlike the other LLMs which had a more modest specificity between 33% and 93% resulting in at least 794 false positive classifications, much greater than the 103 true positive detections (see Table 3).

**Table 3:**
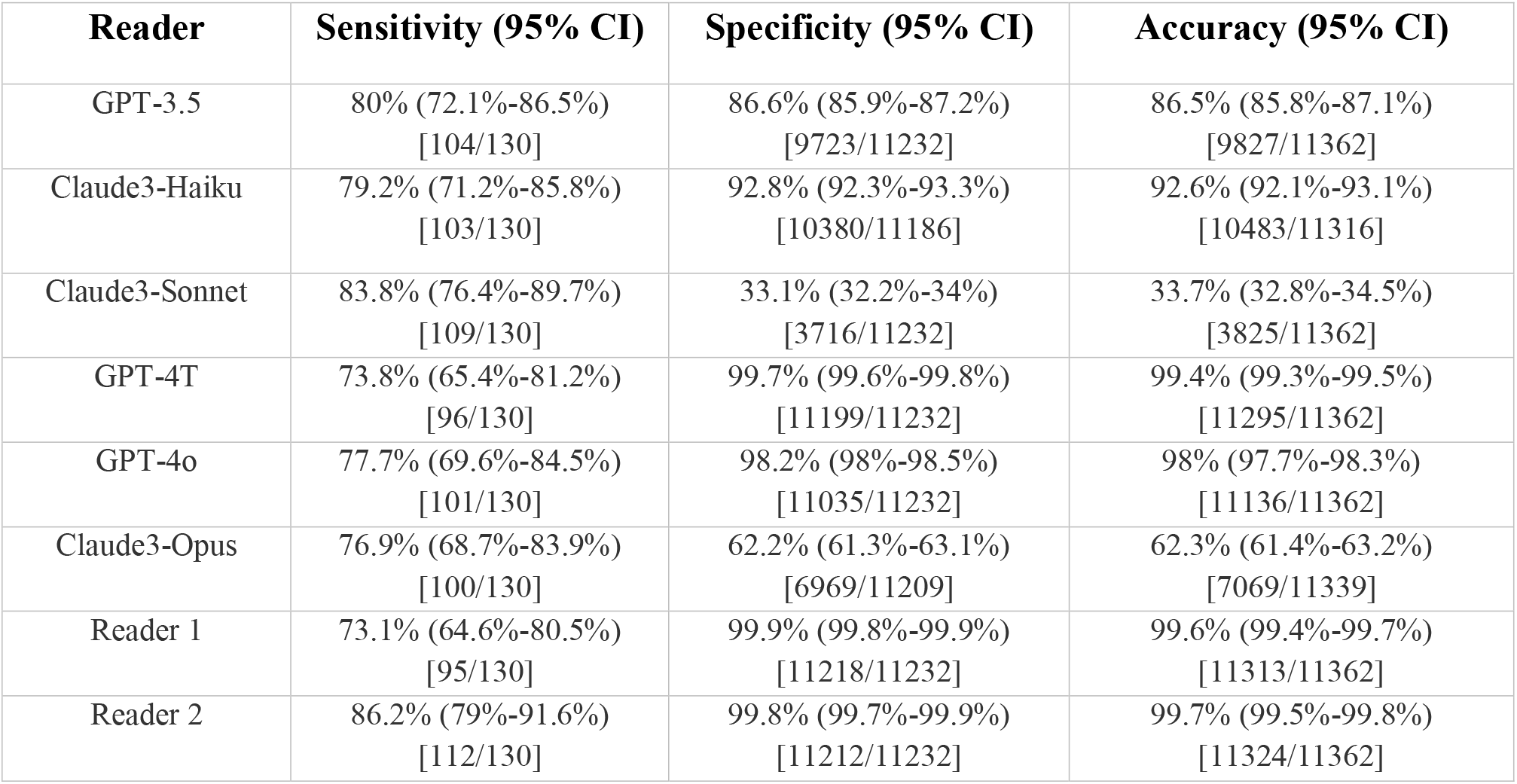
Diagnostic test accuracy of LLMs for specific features of interest detection in Review 2. Human reader performance is shown for context.

| Reader | Sensitivity (95% CI) | Specificity (95% CI) | Accuracy (95% CI) |
| --- | --- | --- | --- |
| GPT-3.5 | 80% (72.1%-86.5%)<br>[104/130] | 86.6% (85.9%-87.2%)<br>[9723/11232] | 86.5% (85.8%-87.1%)<br>[9827/11362] |
| Claude3-Haiku | 79.2% (71.2%-85.8%)<br>[103/130] | 92.8% (92.3%-93.3%)<br>[10380/11186] | 92.6% (92.1%-93.1%)<br>[10483/11316] |
| Claude3-Sonnet | 83.8% (76.4%-89.7%)<br>[109/130] | 33.1% (32.2%-34%)<br>[3716/11232] | 33.7% (32.8%-34.5%)<br>[3825/11362] |
| GPT-4T | 73.8% (65.4%-81.2%)<br>[96/130] | 99.7% (99.6%-99.8%)<br>[11199/11232] | 99.4% (99.3%-99.5%)<br>[11295/11362] |
| GPT-4o | 77.7% (69.6%-84.5%)<br>[101/130] | 98.2% (98%-98.5%)<br>[11035/11232] | 98% (97.7%-98.3%)<br>[11136/11362] |
| Claude3-Opus | 76.9% (68.7%-83.9%)<br>[100/130] | 62.2% (61.3%-63.1%)<br>[6969/11209] | 62.3% (61.4%-63.2%)<br>[7069/11339] |
| Reader 1 | 73.1% (64.6%-80.5%)<br>[95/130] | 99.9% (99.8%-99.9%)<br>[11218/11232] | 99.6% (99.4%-99.7%)<br>[11313/11362] |
| Reader 2 | 86.2% (79%-91.6%)<br>[112/130] | 99.8% (99.7%-99.9%)<br>[11212/11232] | 99.7% (99.5%-99.8%)<br>[11324/11362] |

### Secondary Outcome

#### Automation rate and error rate (Review 1)

The four different scenarios of LLM assistance for abstract screening yielded different automation rates and error rates, depending on the scenario and LLM used (Figure 4). In general, the highest automation rate could be achieved using two high parameter LLMs, however this method would also lead to a high error rate. The highest automation rate, while maintaining a low error rate could be achieved with a single or a combination of two low-parameter models.

**Figure 4:**
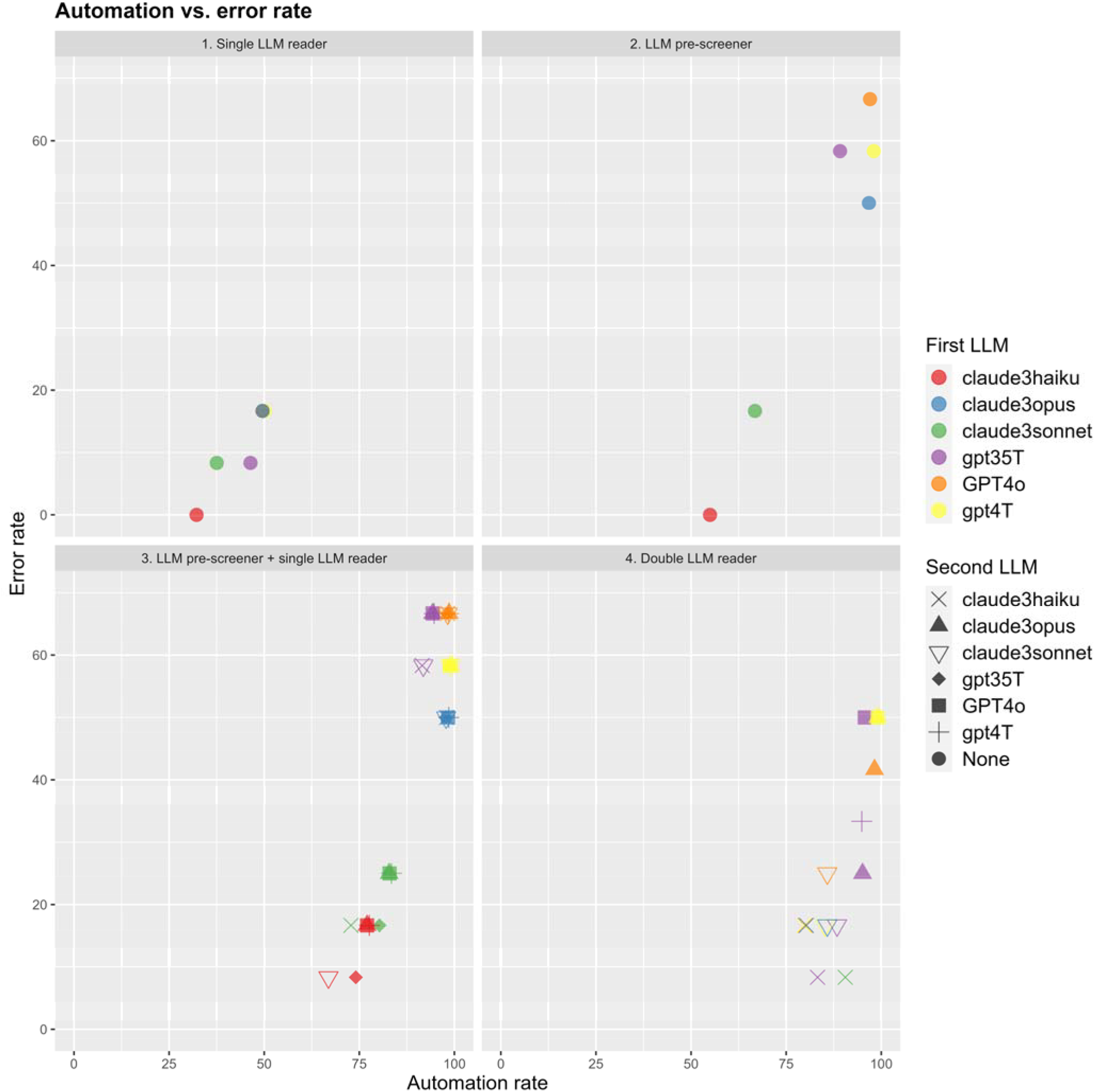
The percentage of overlooked relevant studies (error rate) (y-axis) by the automation rate (x-axis) in four different LLM-assisted abstract screening scenarios. Automation rate refers to the reduction in human abstract screening and arbitration. Multiple data points are overlapping. The # denotes theoretical perfect performance with 100% automation and 0% error rate.

In the single LLM reader scenario (panel 1, Figure 4) we found that using the LLM Claude3-Haiku would reduce the amount of human reads by 32% without errors. The other LLMs would cause abstracts to be overlooked in this scenario, and the highest automation rate would be 50% with GPT-4T, with 16.7% error rate.

In the LLM pre-screen scenario (panel 2, Figure 4) we found the lowest error rate with Claude3-Haiku, which would yield 0% errors, and 55% automation. Highest automation in this scenario would be with GPT-4T, which would lead to 98% automation at a 58% error rate.

In the single LLM reader + LLM pre-screen scenario (panel 3, Figure 4) the lowest error rate was 8% with Claude3-Haiku as pre-screener, and Claude3-Sonnet as reader, which yielded an automation rate of 67%. A similar error rate, but automation rate of 74% was observed with Claude3-Haiku as pre-screener, and GPT-3.5 as reader. The highest automation rate observed was 99% with GPT-4T as pre-screener, and Claude3-Opus as reader, however in this case there was a 58% error rate.

In the double LLM reader scenario (panel 4, Figure 4) the lowest error rate was 8% with Claude3-Haiku and Claude3-Sonnet with an automation rate of 91%. The highest automation rate of 99% was observed with GPT-4T and GPT-4T-o, at 50% error rate.

### Tertiary Outcome

#### Inter- and intra-reader variability

For abstract inclusion in review 1, the inter-reader agreement between human reader 1 and reader 2 was fair (κ=0.25). Moderate inter-reader agreement was observed between the three high parameter models (κ=0.44-0.61), while inter-reader agreement between some of the lower parameter models was poor (κ between 0.03 - 0.19). Agreement between reader 1 and GPT-4o (κ=0.32) and with GPT-4T (κ=0.30) was higher than with reader 2. Reader 2 had higher agreement with Claude3-Sonnet (κ=0.28) and Claude3-Opus (κ=0.27) than with Reader 1.

In review 2 for abstract characterisation, the inter-reader agreement was good in between all LLMs (κ between 0.75 - 0.93). Agreement between readers and models ranged from κ=0.75 (GPT-3.5 vs reader 2) to κ=0.88 (GPT-4T vs reader 1), compared to κ=0.87 between the two readers. For specific features of interest extractions, the agreement between all LLMs was generally low (κ=0.02 and 0.31) with the exception of GPT-4T vs GPT-4o, which had a moderate (κ=0.5) agreement. Agreement between GPT-4T and human readers was moderate to high (κ=0.66 and κ=0.72) while it was poor to moderate (κ=0.01 to 0.44) for the other models. This should be compared to the human inter-reader agreement which was κ=0.64.

Intra-reader agreement was generally high (κ=0.63 to κ>0.99) across models and tasks, except for GPT-3.5 in review 1 (κ=0.51 at temp = 0.8) and GPT-3.5 and Claude3 models in Review 2 (κ=0.50-0.57; table 4). GPT-4o had perfect intra-reader agreement in Review 2, with all outputs identical. In review 1, all models showed lower intra-reader agreement for abstract inclusion at higher temperatures.

**Table 4:** Intra-reader agreement of the LLMs. Numbers display Cohens Kappa value calculated by comparing two runs with the LLMs.

| Intra-reader agreement of the LLMs |  |  |  |  |
| --- | --- | --- | --- | --- |
|  | Inclusion in Systematic Review, temperature = 0.2 | Inclusion in Systematic Review, temperature = 0.8 | Abstract Characterisation in Scoping Review, temperature = 1.0 | Specific Feature of Interest in Scoping Review, temperature = 1.0 |
| GPT-3.5 | 0.73 | 0.51 | 0.89 | 0.50 |
| Claude3-Haiku | 0.95 | 0.81 | 0.92 | 0.57 |
| Claude3-Sonnet | 0.95 | 0.76 | 0.94 | 0.55 |
| GPT-4T | 0.81 | 0.63 | 0.98 | 0.85 |
| GPT-4o | 0.96 | 0.76 | 1.0 | 1.0 |
| Claude3-Opus | 0.91 | 0.87 | 0.93 | 0.56 |

**Table 5:** GLMM of model accuracy for correct abstract inclusion in review 1.^*^OR of accurate abstract inclusion compared to GPT-4o. The 11808 abstract decisions stem from two runs at temp 0.2 and two runs at temp 0.8 with 492 abstracts and six LLMs using the inclusion criteria prompt strategy.

| GLMM of model accuracy for correct abstract inclusion in review 1 |  |  |
| --- | --- | --- |
| LLM | OR (95%CI) | p-value |
| GPT-3.5* | 0.041 (0.029-0.058) | <0.001 |
| Claude3-Haiku* | 0.001 (0.000-0.001) | <0.001 |
| Claude3-Sonnet* | 0.109 (0.077-0.155) | <0.001 |
| GPT-4T* | 1.066 (0.711-1.598) | 0.8 |
| Claude3-Opus* | 1.043 (0.697-1.562) | 0.8 |
| Temperature (0.8 vs. 0.2) | 1.104 (0.939-1.299) | 0.2 |

#### Other prompt strategies

To test the prompt dependency of our results in review 1, we tried two different prompt strategies. First, we tried to screen the abstracts with only inclusion criteria, and no exclusion criteria. This strategy increased the accuracy of Claude3-Sonnet, but interestingly it led to a decrease in the accuracy of GPT-3.5 and Claude3-Haiku. It had minimal effect on the high parameter models. Afterwards we prompted the LLMs to output a confidence score from 0-100 of how well the inclusion and exclusion criteria were met, to calculate ROC-AUC, and specificities fixed at human reader 1 and 2’s sensitivities (Figure 5). Here we found that no LLM was on level with the human specificity, however GPT-4T achieved a 90% specificity at reader 1 equivalent sensitivity and 94% specificity at reader 2 equivalent sensitivity. All numbers are available in the supplementary materials.

**Figure 5:**
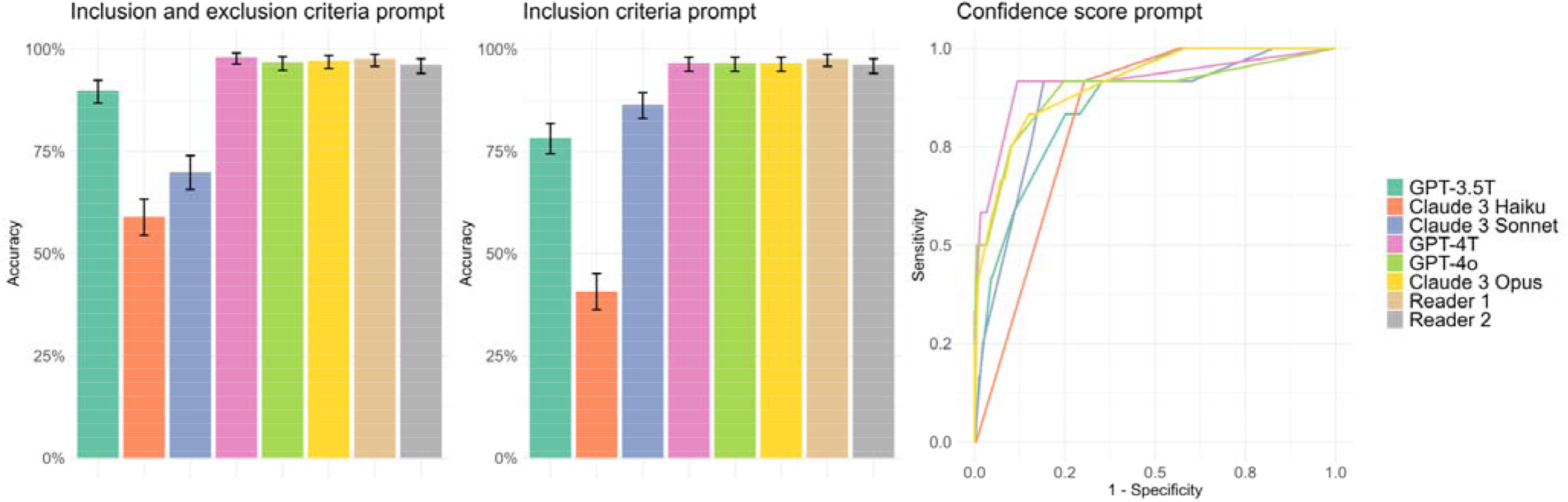
Accuracy and receiver operating curves for each LLM in correct abstract inclusion in the systematic review (review 1). In the confidence score prompt, ROC-AUCs were the following: 0.867 (GPT-3.5), 0.822 (Claude3-Haiku), 0.864 (Claude3-Sonnet), 0.916 (GPT-4T), 0.889 (GPT-4o), 0.909 (Claude3-Opus).

### Association between accuracy, LLM, and temperature

With a generalised linear mixed model (GLMM) of 11808 abstract inclusion decisions in review 1, we found no differences between the accuracy of GPT-4T and GPT-4o, and Claude3-Opus and GPT-4o, when we adjusted for different temperature levels. We also found that model temperature was not independently associated with the accuracy of the LLMs (table 4).

### False negative analysis of LLMs for review 1

We conducted a qualitative analysis of the 12 abstracts included in review 1 by the human reference, to understand the disagreement by the different readers and LLMs with the primary prompt. Overall GPT-4T, GPT-4o, and Claude3-Opus were very strict in the evaluation of the abstracts, but committed few errors in the text understanding. Meanwhile, the human readers were a bit inaccurate with regards to the comparator criteria, especially. This may explain the low sensitivity observed in those LLMs. Meanwhile it seems that GPT-3.5 commits more errors related to understanding the text. The full qualitative analysis is available in the supplementary materials. If we excluded the seven studies where human inclusion could be considered wrong, the sensitivities of correct abstract inclusion were the following: reader 1 (100%), reader 2 (80%), GPT-3.5 (40%), GPT-4T (80%), GPT-4o (80%), Claude3-Opus (80%), Claude3-Sonnet (100%), Claude3-Haiku (100%). The combination of GPT-4T and GPT-4o as reader 1 and 2, with a human reader for disagreements, would include all relevant abstracts, and reduce the number of manual reads by 99%. The same could be achieved with GPT-4T and Claude3-Opus, but at a higher cost.

### Post-hoc extrapolation to full systematic review

Extrapolating to the full systematic review, with 6.3% human disagreements the combination of GPT-4T and GPT-4o would mean a reduction in the number of human abstract reads from 8859^*^2+558 = 18276 abstract reads to 183 arbitration reads. Abstract screening has been reported to average 1.7 minutes per abstract in systematic reviews^18^. In time, this would equal to a reduction from 18276^*^1.7 min/abstract = 518 hours of work to 5.2 hours of work. In cost, the full abstract screening with two human readers and a third for arbitrations would cost 518 hours^*^40.9 USD/hour = 21,186 USD. Meanwhile, the LLM-assisted abstract screening with GPT-4T and GPT-4o would cost 8859^*^0.01011 USD = 89.5 USD, and 8859^*^0.00508 USD = 45 USD in API fees and 5.2^*^40.9 USD = 213 USD in human arbitration hour salary for a total of 347.5 USD, which is 60 times cheaper than the conventional two human readers method.

## Discussion

We conducted a diagnostic test accuracy study of six commercially available LLMs for abstract characterisation, screening abstracts for inclusion and detection of specific features in systematic and scoping reviews.

For abstract characterisation all LLM’s could determine study type, language and study participants with near perfect accuracy. Only higher parameter models (GPT-4o, GPT-4T and Claude3-Opus) were able to reliably determine if participants had a specific disease. For features of interest detection only GPT-4T and GPT-4o achieved very high and close to human accuracy, while the other LLMs suffered from a large number of false positive findings.

For abstract screening for inclusion according to predefined PICO criteria GPT-4o, GPT-4T and Claude3-Opus achieved accuracy, which was on level with the human readers but at only a moderate sensitivity while lower parameter models Claude3-Haiku and Claude3-Sonnet achieved a high sensitivity but at only moderate specificity.

Comparing to existing literature, a recent study on title and abstract screening using LLMs adopted a scale from 1 to 5 on how relevant a study was for the systematic review^11^. The study used previously published systematic review datasets, which were analysed with the LLMs FlanT5, OpenHermes 2.5, Mixtral and Platypus 2. In their study, they found high sensitivities (82-100%), and low specificities (13%-75%), similar to the performance of the lower parameter models in our study.

Another study tested GPT-3.5 and GPT-4T LLMs on previously published review datasets from the literature^12^. Similarly to our study, they informed the LLMs of the title, abstract, inclusion and exclusion criteria, and then gave instructions to include or exclude. They found an overall accuracy of 91%, sensitivity of 76%, and specificity of 91% for the GPT-models, but did not report diagnostic performance individually for the two LLMs. This makes it difficult to compare to our results, as we observed that the sensitivity-specificity cutoff was very different for the different LLMs. Interestingly they found that the sensitivities and specificities displayed large heterogeneity between the 6 different datasets they tested on. This may be attributable to the prompting strategies, inclusion/exclusion criteria, but also to the ground truth of the human screening. They found a higher agreement in the 6 studies between human readers than we did in ours (κ=0.46 vs. κ=0.25), suggesting that our systematic review criteria may be difficult for human readers to agree on.

A third recent study evaluated GPT-4 (unknown version) on screening abstracts and full text studies^13^. They found lower accuracy than us for title and abstract screening (67%-88%), but close to human performance for full text screening (54%-96%). We were unable to find studies on abstract characterisation, feature of interest detection with LLMs, model temperature or different implementation scenarios, highlighting our study’s contribution to the field.

Overall results from this study corroborate the existing evidence that LLMs can be used with high accuracy for select PICOs based abstract screening, with frontier models performing at higher specificity and lower sensitivity. It therefore seems prudent, to manually read a subset of abstracts to establish the sensitivity of an LLM for a given PICOs, if LLMs are to be used in systematic reviews.

Our study found, that a double LLM abstract screening with human arbitration in case of disagreement achieved the highest automation rate of up to 91%, with a low error rate of 8%. When no abstracts are to be overlooked during screening, we found that an abstract pre-screening with a high sensitivity LLM (Claude3-Haiku) was able to reduce the human workload by more than 50%.

Using 1.7 minutes per title and abstract screening, we can estimate our full systematic review abstract screening to take 518 hours to complete (65 8-hour workdays), which would be reduced to 49.2 hours of work with the double LLM-reader scenario with Claude 3.5 Sonnet and Haiku. The potential for time-saving with LLM assistance is in other words very great.

Future studies should explore if LLMs can assist in other parts of the review process, such as full-text screening, risk of bias assessment and extraction of outcome variables. In addition, future studies may experiment in greater depth with newer generations of LLMs, different prompt constructions or explore chain-of-thought prompting^19–21^. Chain-of-thought prompting has been associated with better performance in tasks where LLMs otherwise did not outperform the average human rater^19^.

Potential problems with using LLMs for reviews are that some LLMs can confabulate and hallucinate^22,23^. Another potential problem is the risk of bias in LLMs^24,25^, where for example a bias against studies from certain countries or patients would be unwanted. Finally LLM guardrails, a common public safety feature which prevent return of answers addressing sensitive topics, could negatively impact some research subjects (e.g. suicide prevention studies in psychiatry).

There are limitations to our study. Firstly, we only included commercially available LLMs from two different companies. Secondly, LLMs are prone to confabulations and hallucinations. Since we limited the results of the LLM to a structured output, we were not able to determine if incorrect results were due to hallucinations. Thirdly, the estimated human reader performance might be overestimated, due to confounding bias, where the reader and reference are not independent. In two of the abstract characterisation prompts we found a low agreement between two of the readers regarding organ and the modality used and low accuracy of the LLMs used. This may have been a result of too vaguely defined abstract characteristics. Fourthly, in the primary outcome we forced a strict classification (include or exclude) unto the LLMs, but during exploratory investigation found that adding confidence estimates to the LLM-output actually resulted in a more favourable performance. Finally, the number of abstracts included in the systematic review was large (500), but only a small portion (12 abstracts) were actually included by the reference standard, meaning that the dataset was imbalanced and confidence intervals of sensitivity are wide.

Our study adds to existing literature by demonstrating that LLMs can be used for scoping reviews and by developing different LLM-assisted screening scenarios. A strength of our study approach was that we tested the LLMs on new, unpublished, systematic and scoping reviews. The LLMs could therefore not draw on previously published reviews, which could have been part of the training data for the LLMs.

In conclusion, we demonstrated that abstract characterisation and specific feature of interest detection with LLMs is feasible and accurate with GPT-4o and GPT-4T. The majority of abstract screenings for systematic review could be automated with a double LLM-reader scenario at a low error rate.

## Data Availability

The data used in this study are available on request from the corresponding author.

## References

1. Hardi AC, Fowler SA. Evidence-Based Medicine and Systematic Review Services at Becker Medical Library. Mo Med 2014;111:416–8.

2. Government of Canada CI of HR. A Guide to Knowledge Synthesis - CIHR. 2010 Mar 25. [Epub ahead of print].

3. Page MJ, McKenzie JE, Bossuyt PM, et al. The PRISMA 2020 statement: an updated guideline for reporting systematic reviews. BMJ 2021;372:71.

4. Belur J, Tompson L, Thornton A, et al. Interrater Reliability in Systematic Review Methodology: Exploring Variation in Coder Decision-Making. Sociol Methods Res 2021;50:837–65.

5. Hanegraaf P, Wondimu A, Mosselman JJ, et al. Inter-reviewer reliability of human literature reviewing and implications for the introduction of machine-assisted systematic reviews: a mixed-methods review. BMJ Open 2024;14:e076912.

6. Rousson V, Gasser T, Seifert B. Assessing intrarater, interrater and test-retest reliability of continuous measurements. Stat Med 2002;21:3431–46.

7. McGowan J, Sampson M. Systematic reviews need systematic searchers. J Med Libr Assoc 2005;93:74– 80.

8. Borah R, Brown AW, Capers PL, et al. Analysis of the time and workers needed to conduct systematic reviews of medical interventions using data from the PROSPERO registry. BMJ Open 2017;7:e012545.

9. PubMed NCBI.

10. Qureshi R, Shaughnessy D, Gill KAR, et al. Are ChatGPT and large language models “the answer” to bringing us closer to systematic review automation? Syst Rev 2023;12:72.

11. Dennstädt F, Zink J, Putora PM, et al. Title and abstract screening for literature reviews using large language models: an exploratory study in the biomedical domain. Syst Rev 2024;13:158.

12. Guo E, Gupta M, Deng J, et al. Automated Paper Screening for Clinical Reviews Using Large Language Models: Data Analysis Study. J Med Internet Res 2024;26:e48996.

13. Khraisha Q, Put S, Kappenberg J, et al. Can large language models replace humans in systematic reviews? Evaluating GPT-4’s efficacy in screening and extracting data from peer-reviewed and grey literature in multiple languages. Res Synth Methods 10.1002/jrsm.1715.

14. Ambalavanan AK, Devarakonda MV. Using the contextual language model BERT for multi-criteria classification of scientific articles. J Biomed Inform 2020;112:103578.

15. Issaiy M, Ghanaati H, Kolahi S, et al. Methodological insights into ChatGPT’s screening performance in systematic reviews. BMC Med Res Methodol 2024;24:78.

16. Cohen JF, Korevaar DA, Altman DG, et al. STARD 2015 guidelines for reporting diagnostic accuracy studies: explanation and elaboration. BMJ Open 2016;6:e012799.

17. Friedli I, Baid-Agrawal S, Unwin R, et al. Magnetic Resonance Imaging in Clinical Trials of Diabetic Kidney Disease. J Clin Med 2023;12:4625.

18. Citation screening in systematic reviews: two approaches, two authors and time taken (SWAR-1 (Study Within A Review 1)) Cochrane Colloquium Abstracts.

19. Wei J, Wang X, Schuurmans D, et al. Chain-of-Thought Prompting Elicits Reasoning in Large Language Models. Adv Neural Inf Process Syst 2022;35:24824–37.

20. Zhang Z, Zhang A, Li M, et al. Automatic Chain of Thought Prompting in Large Language Models. 10.48550/arXiv.2210.03493.

21. Suzgun M, Scales N, Schärli N, et al. Challenging BIG-Bench Tasks and Whether Chain-of-Thought Can Solve Them. 10.48550/arXiv.2210.09261.

22. Farquhar S, Kossen J, Kuhn L, et al. Detecting hallucinations in large language models using semantic entropy. Nature 2024;630:625–30.

23. Gunjal A, Yin J, Bas E. Detecting and Preventing Hallucinations in Large Vision Language Models. Proc AAAI Conf Artif Intell 2024;38:18135–43.

24. Kotek H, Dockum R, Sun D. Gender bias and stereotypes in Large Language Models. In: Proceedings of The ACM Collective Intelligence Conference. CI ‘23. New York, NY, USA: Association for Computing Machinery; 2023:12–24.

25. Navigli R, Conia S, Ross B. Biases in Large Language Models: Origins, Inventory, and Discussion. J Data Inf Qual 2023;15:10:1-10:21.

