## supplementary materials for "Large language models for abstract screening in systematic- and scoping reviews: A diagnostic test accuracy study"

### S1: STARD-2015-checklist

| **Section and Topic** | **No** | **Item** | **Check** |
| --- | --- | --- | --- |
| Title or Abstract |  |  |  |
|  | 1 | Identification as a study of diagnostic accuracy using at least one measure of accuracy (such as sensitivity, specificity, predictive values, or AUC) | x |
| Abstract |  |  |  |
|  | 2 | Structured summary of study design, methods, results, and conclusions (for specific guidance, see STARD for Abstracts) | x |
| Introduction |  |  |  |
|  | 3 | Scientific and clinical background, including the intended use and clinical role of the index test | x |
|  | 4 | Study objectives and hypotheses | x |
| Methods |  |  |  |
| Study design | 5 | Whether data collection was planned before the index test and reference standard were performed (prospective study) or after (retrospective study) | x |
| Participants |  |  |  |
|  | 6 | Eligibility criteria | x |
|  | 7 | On what basis potentially eligible participants were identified (such as symptoms, results from previous tests, inclusion in registry) | x |
|  | 8 | Where and when potentially eligible participants were identified (setting, location, and dates) | x |
|  | 9 | Whether participants formed a consecutive, random, or convenience series | x |
| Test methods |  |  |  |
|  | 10a | Index test, in sufficient detail to allow replication | x |
|  | 10b | Reference standard, in sufficient detail to allow replication | x |
|  | 11a | Rationale for choosing the reference standard (if alternatives exist) | x |
|  | 12a | Definition of and rationale for test positivity cut-offs or result categories of the index test, distinguishing prespecified from exploratory | x |
|  | 12b | Definition of and rationale for test positivity cut-offs or result categories of the reference standard, distinguishing prespecified from exploratory | x |
|  | 13a | Whether clinical information and reference standard results were available to the performers or readers of the index test | x |
|  | 13b | Whether clinical information and index test results were available to the assessors of the reference standard | x |
| Analysis |  |  |  |
|  | 14 | Methods for estimating or comparing measures of diagnostic accuracy | x |
|  | 15 | How indeterminate index test or reference standard results were handled | x |
|  | 16 | How missing data on the index test and reference standard were handled | x |
|  | 17 | Any analyses of variability in diagnostic accuracy, distinguishing prespecified from exploratory | % |
|  | 18 | Intended sample size and how it was determined | % |
| Results |  |  |  |
| Participants | 19 | Flow of participants, using a diagram | NA |
|  | 20 | Baseline demographic and clinical characteristics of participants | NA |
|  | 21a | Distribution of severity of disease in those with the target condition | NA |
|  | 21b | Distribution of alternative diagnoses in those without the target condition | NA |
| Test results |  |  |  |
|  | 22 | Time interval and any clinical interventions between index test and reference standard | NA |
|  | 23 | Cross-tabulation of the index test results (or their distribution) by the results of the reference standard | x |
|  | 24 | Estimates of diagnostic accuracy and their precision (such as 95% CIs) | x |
|  | 25 | Any adverse events from performing the index test or the reference standard | NA |
| Discussion |  |  |  |
|  | 26 | Study limitations, including sources of potential bias, statistical uncertainty, and generalizability | x |
|  | 27 | Implications for practice, including the intended use and clinical role of the index test | x |
| Other Information |  |  |  |
|  | 28 | Registration number and name of registry | % |
|  | 29 | Where the full study protocol can be accessed | % |
|  | 30 | Sources of funding and other support; role of funders | x |

### S2: Pubmed-search-strategy

###### Review 1

search_string_1 = '("Central Nervous System"[Mesh]) OR (brain*[All Fields]) OR (cerebr*[All Fields])'
search_string_2 = '("Diagnostic Imaging"[Mesh] OR CT[All Fields]) OR (MRI[All Fields])'
search_string_3 = '("Deep Learning"[Mesh]) OR ("Neural Networks, Computer"[Mesh]) OR (Neural network*[All Fields]) OR (Convolutional network*[All Fields]) OR (Deep learn*[All Fields]) OR (Artificial Intelligence*[All Fields])'
date_range = '("2017/01/01"[PDAT] : "2024/04/23"[PDAT])'
reviews = '(review[Publication Type])'

combined_search_string = f"({search_string_1}) AND ({search_string_2}) AND ({search_string_3}) AND ({date_range}) NOT ({reviews})"

###### Review 2

("diabetes mellitus"[MeSH Terms] OR ("diabetes"[All Fields] AND "mellitus"[All Fields]) OR "diabetes mellitus"[All Fields])

AND

("kidney"[MeSH Terms] OR "kidney"[All Fields] OR "kidneys"[All Fields] OR "kidney s"[All Fields])

NOT "Review"[PT] NOT "Case Reports"[PT] NOT "Comment"[PT] NOT "Editorial"[PT] NOT "Systematic Review"[PT]

AND "English"[LA]

AND "Diagnostic Imaging"[MeSH Terms]'

### S3: LLM-prompts

###### Review 1

**Prompt strategy 1: PICO-inclusion and exclusion prompt**

"content": "*You are a medical researcher analyzing abstracts for a systematic review. Answer concisely in JSON format with only the following keys: population_include, population_exclude, intervention_include, intervention_exclude, comparator_include, comparator_exclude, outcome_include, outcome_exclude, study_characteristics_include, study_characteristics_exclude, decision.*"

llm_prompt = '*Please assess the title and abstract based on the following criteria for inclusion in a systematic review and provide a final decision:*

- *population_include: Adult patients with suspected neurological disease. [yes/no/NA]*
- *population_exclude: Pediatric studies, animal studies.[yes/no/NA]*
- *intervention_include: Analysis of brain scan with a deep learning or neural network algorithm/tool.[yes/no/NA]*
- *intervention_exclude: Other modalities such as EEG, MEG, and ultrasound.[yes/no/NA]*
- *comparator_include: Manual analysis of brain scan (radiology report or expert reader).[yes/no/NA]*
- *comparator_exclude: If no reference test or ground truth is mentioned.[yes/no/NA]*
- *outcome_include: Diagnostic test accuracy of the intervention in an external cohort.[yes/no/NA]*
- *outcome_exclude: If only internal testing is performed (for example by random data splitting or cross-validation).[yes/no/NA]*
- *study_characteristics_include: Diagnostic test accuracy studies mentioning sensitivity, specificity, accuracy, or AUC for diagnostic performance. [yes/no/NA]*
- *study_characteristics_exclude: Non-diagnostic test accuracy studies.Only other outcomes, such as DICE score. [yes/no/NA]*

*decision: Include if all the inclusion criteria are met and no exclusion criteria are met. Exclude if any exclusion criteria are met. Uncertain if no exclusion criteria are met but one of the inclusion criteria is unclear in the abstract, but the other inclusion criteria are met. [include/exclude/uncertain]*

*Title and abstract:*'

For prompt 1 we ran the analysis one time at temperature 0.2.

**Prompt strategy 2: PICO-inclusion criteria only prompt**

"content": "*You are a medical researcher analyzing abstracts for a systematic review. Answer in JSON format with the following keys: population, intervention, control, outcome, decision.*"

llm_prompt = '*Please assess the title and abstract based on the following criteria for inclusion in a systematic review: population: Does the study examine adults with suspected neurological disease(s)? [yes/no/NA]. intervention: Does the study use neural networks or deep learning for brain scan analysis? [yes/no/NA]. control: Does the study compare these methods against a standard reference (radiological report or expert readers)? [yes/no/NA]. outcome: Does it measure the diagnostic accuracy of the intervention using an external cohort for validation? [yes/no/NA]. decision: Include if all the above criteria are met, exclude if any criteria are not met, uncertain if information is insufficient [include/exclude/uncertain]. Title and abstract:*'

For prompt 2, we repeated the LLM analysis four times at temperature 0.2 and four times at temperature 0.8 to evaluate consistency in the outputs from the LLMs and the effect of different LLM temperature.

**Prompt strategy 3: decision score 0-100:**

"content": "*You are a medical researcher analyzing abstracts for a systematic review. Answer concisely in JSON format with only the following key: decision_score.*"

llm_prompt = '*Please assess the title and abstract based on the following criteria for inclusion for full-text review in a systematic review and provide a final decision score from 0 to 100, where 0 means certain exclusion and 100 means certain inclusion. Exclusion criteria should lower the score, while inclusion criteria should increase the score. A score of 100 should only be given if all inclusion criteria are met and no exclusion criteria are met.*

*Population include: Adult patients with suspected neurological disease.*

*Population exclude: Pediatric studies, Animal studies.*

*Intervention include: Analysis of brain scan with a deep learning or neural network algorithm/tool.*

*Intervention exclude: Other modalities such as EEG, MEG, and ultrasound.*

*Comparator include: Manual analysis of brain scan (radiology report or expert reader).*

*Comparator exclude: If no reference test or ground truth is mentioned.*

*Outcome include: Diagnostic test accuracy of the intervention in an external cohort.*

*Outcome exclude: If there is only internal testing performed (for example by random data splitting or cross-validation).*

*Study characteristics include: Diagnostic test accuracy studies mentioning sensitivity, specificity, accuracy, or AUC for diagnostic performance.*

*Study characteristics exclude: Non-diagnostic test accuracy studies. Only other outcomes, such as DICE score. Title and abstract:*'

For prompt 3, we ran the LLM analysis one time at temperature 0.2.

###### Review 2

content = “*You are a medical researcher analyzing abstracts for a systematic review. You are designed to output JSON.*” + [Abstract Title] + [Abstract Text]

llm_prompt = *“Please answer the following questions with one of the indicated responses as concisely as you can.*

- *study: Is the abstract for a review, clinical study, animal study, case report, or something else? [Review/Clinical Study/Animal Study/Case Report/Other/NA]*
- *english: Is the abstract written in English? [Yes/No/NA]*
- *human: Is this a study with human participants, animals or something else? [Humans/Animals/Other/NA]*
- *dm: Does the study have patients with diabetes mellitus as its primary focus? [Yes/No/NA]*
- *modality: Does this study describe the kidney using CT, MRI, Ultrasound, or other radiologic modalities? [CT/MRI/Ultrasound/Other Radiologic Modality/Multiple Modalities/No Radiologic Modality/NA]*
- *organ: Does the study appear to describe changes in the kidney, in other organs or both? [Kidney/Other Organs/Both/NA]*
- *post-contrast kidney injury: Is this a study on post-contrast kidney injury? [Yes/No/Equivocal/NA]*
- *BOLD: Was blood oxygenation level-dependent (BOLD) apparent relaxation rates measured? [Yes/No/NA]*
- *adc: Was apparent diffusion coefficient (ADC) measured? [Yes/No/NA]*
- *t1_relax: Was apparent relaxation rate of T1 weighted images measured? [Yes/No/NA]*
- *perirenal fat: Was perirenal fat measured? [Yes/No/NA]*
- *sinus fat: Was renal sinus fat measured? [Yes/No/NA]*
- *parenchymal fat: Was renal parenchymal fat measured? [Yes/No/NA]*
- *parenchymal volume: Was kidney parenchymal volume measured? [Yes/No/NA]*
- *kidney volume: Was kidney volume measured? [Yes/No/NA]*
- *kidney length: Was kidney length measured? [Yes/No/NA]*
- *kidney size: Was kidney size measured? [Yes/No/NA]*
- *parenchymal heterogeneity: Was parenchymal heterogeneity assessed? [Yes/No/NA]*
- *parenchymal iodine enhancement: Was parenchymal iodine enhancement measured? [Yes/No/NA]*
- *parenchymal thickness: Was parenchymal or cortical thickness measured? [Yes/No/NA]*
- *renal blood flow: Was renal blood flow measured? [Yes/No/NA]*
- *systolic velocity: Was peak systolic velocity measured? [Yes/No/NA]*
- *diastolic velocity: Was end diastolic velocity measured? [Yes/No/NA]*
- *resistive index: Was artery resistive index measured? [Yes/No/NA]*
- *global perfusion: Was global renal perfusion measured? [Yes/No/NA]*
- *global attenuation: Was global renal CT attenuation (as stated in HU values) measured? [Yes/No/NA]*
- *iodine arterial: Was iodine concentration in one or more renal arteries measured? [Yes/No/NA]*
- *iodine venous: Was iodine concentration in one or more renal veins measured? [Yes/No/NA]*
- *focal attenuation: Was focal CT attenuation (as stated in HU values) measured? [Yes/No/NA]*
- *focal iodine enhancement: Was focal iodine enhancement measured? [Yes/No/NA]”*

For the scoping review all analysis with the LLM was performed two times for each model, and at temperature of 1.0.

### S4: Diagnostic test accuracy supplementary tables

###### Review 1: Abstract inclusion with different prompt strategies

| **Prompt strategy two - inclusion criteria only** | | | |
| --- | --- | --- | --- |
| **Reader** | **Sensitivity (95% CI)** | **Specificity (95% CI)** | **Accuracy (95% CI)** |
| GPT-3.5 | 0.67 (0.35-0.9) [8/12] | 0.79 (0.75-0.82) [377/480] | 0.78 (0.74-0.82) [385/492] |
| Claude3-Haiku | 1 (0.74-1) [12/12] | 0.39 (0.35-0.44) [188/480] | 0.41 (0.36-0.45) [200/492] |
| Claude3-Sonnet | 0.75 (0.43-0.95) [9/12] | 0.87 (0.83-0.9) [416/480] | 0.86 (0.83-0.89) [425/492] |
| GPT-4T | 0.25 (0.05-0.57) [3/12] | 0.98 (0.97-0.99) [472/480] | 0.97 (0.95-0.98) [475/492] |
| GPT-4o | 0.42 (0.15-0.72) [5/12] | 0.98 (0.96-0.99) [470/480] | 0.97 (0.95-0.98) [475/492] |
| Claude3-Opus | 0.25 (0.05-0.57) [3/12] | 0.98 (0.97-0.99) [472/480] | 0.97 (0.95-0.98) [475/492] |
| Reader 2 | 0.67 (0.35-0.9) [8/12] | 0.97 (0.95-0.98) [465/480] | 0.96 (0.94-0.98) [473/492] |
| Reader 1 | 0.83 (0.52-0.98) [10/12] | 0.98 (0.96-0.99) [470/480] | 0.98 (0.96-0.99) [480/492] |

| **Prompt strategy three - confidence score from 0-100** | | | | | | | | | |
| --- | --- | --- | --- | --- | --- | --- | --- | --- | --- |
|  |  | **Reader 1** | **Reader 2** | **GPT-4o** | **GPT-4T** | **GPT-3.5** | **Claude3-Opus** | **Claude3-Sonnet** | **Claude3-Haiku** |
| **PICO prompt with confidence score 0-100** | **AUC** | **-** | **-** | 0.889 | 0.916 | 0.867 | 0.909 | 0.864 | 0.822 |
|  | **Specificity at 100% sensitivity** | **-** | **-** | 0.00 | 0.00 | 0.43 | 0.42 | 0.17 | 0.43 |
|  | **Specificity at reader 1 sensitivity** | **0.98 (0.96-0.99) [470/480]** | **-** | 0.83 | 0.90 | 0.75 | 0.85 | 0.83 | 0.72 |
|  | **Specificity at reader 2 sensitivity** | **-** | **0.97 (0.95-0.98) [465/480]** | 0.92 | 0.94 | 0.84 | 0.92 | 0.89 | 0.78 |

###### Review 2: Abstract characteristics

| Supplemental Table | | | | | | | |
| --- | --- | --- | --- | --- | --- | --- | --- |
| Accuracy (95% Conf Int) for Abstract Characteristics extraction | | | | | | | |
| Reader | Diabetes | Language | Human | Modality | Organ | PC-AKI | Study Type |
| GPT-3.5 | 74.5% (70.4%-78.3%) [368/494] | 100% (99.3%-100%) [494/494] | 97% (95%-98.3%) [479/494] | 65% (60.6%-69.2%) [321/494] | 47.4% (42.9%-51.9%) [234/494] | 65.2% (60.8%-69.4%) [322/494] | 96.2% (94.1%-97.7%) [475/494] |
| GPT-4o | 95.1% (92.9%-96.9%) [470/494] | 100% (99.3%-100%) [494/494] | 97.4% (95.5%-98.6%) [481/494] | 67% (62.7%-71.1%) [331/494] | 58.7% (54.2%-63.1%) [290/494] | 98.8% (97.4%-99.6%) [488/494] | 94.9% (92.6%-96.7%) [469/494] |
| GPT-4T | 95.3% (93.1%-97%) [471/494] | 100% (99.3%-100%) [494/494] | 98% (96.3%-99%) [484/494] | 64.2% (59.8%-68.4%) [317/494] | 64% (59.6%-68.2%) [316/494] | 99% (97.7%-99.7%) [489/494] | 96.8% (94.8%-98.1%) [478/494] |
| Claude3-Haiku | 79.1% (75.2%-82.6%) [389/492] | 100% (99.3%-100%) [492/492] | 97.6% (95.8%-98.7%) [480/492] | 63.4% (59%-67.7%) [312/492] | 56.9% (52.4%-61.3%) [280/492] | 90.7% (87.7%-93.1%) [446/492] | 95.3% (93.1%-97%) [469/492] |
| Claude3-Opus | 94.7% (92.4%-96.5%) [467/493] | 100% (99.3%-100%) [493/493] | 97.6% (95.8%-98.7%) [481/493] | 67.5% (63.2%-71.7%) [333/493] | 65.1% (60.7%-69.3%) [321/493] | 66.7% (62.4%-70.9%) [329/493] | 96.1% (94%-97.7%) [474/493] |
| Reader 1 | 95.3% (93.1%-97%) [471/494] | 100% (99.3%-100%) [494/494] | 97.4% (95.5%-98.6%) [481/494] | 95.1% (92.9%-96.9%) [470/494] | 77.1% (73.2%-80.8%) [381/494] | 98% (96.3%-99%) [484/494] | 96.8% (94.8%-98.1%) [478/494] |
| Reader 2 | 96.6% (94.5%-98%) [477/494] | 100% (99.3%-100%) [494/494] | 99.2% (97.9%-99.8%) [490/494] | 71.5% (67.3%-75.4%) [353/494] | 94.1% (91.7%-96%) [465/494] | 100% (99.3%-100%) [494/494] | 98.4% (96.8%-99.3%) [486/494] |
| Claude3-Sonnet | 91.5% (88.7%-93.8%) [452/494] | 100% (99.3%-100%) [494/494] | 98.2% (96.6%-99.2%) [485/494] | 65.4% (61%-69.6%) [323/494] | 66.6% (62.2%-70.7%) [329/494] | 78.1% (74.2%-81.7%) [386/494] | 96.6% (94.5%-98%) [477/494] |

### S5: Inter-reader variability supplementary tables

###### Review 1: Inter-reader variability between the reference and LLMs for review 1

|  | **reader_1** | **reader_2** | **reference** | **GPT-3.5** | **GPT-4T** | **GPT-4o** | **claude3opus** | **claude3sonnet** | **claude3haiku** |
| --- | --- | --- | --- | --- | --- | --- | --- | --- | --- |
| **reader_1** | - | 0.25 | 0.61 | 0.12 | 0.3 | 0.32 | 0.24 | 0.21 | 0.05 |
| **reader_2** | 0.25 | - | 0.44 | 0.11 | 0.15 | 0.23 | 0.27 | 0.28 | 0.05 |
| **reference** | 0.61 | 0.44 | - | 0.09 | 0.24 | 0.35 | 0.24 | 0.18 | 0.03 |
| **GPT-3.5** | 0.12 | 0.11 | 0.09 | - | 0.09 | 0.13 | 0.08 | 0.38 | 0.28 |
| **GPT-4T** | 0.3 | 0.15 | 0.24 | 0.09 | - | 0.61 | 0.44 | 0.18 | 0.03 |
| **GPT-4o** | 0.32 | 0.23 | 0.35 | 0.13 | 0.61 | - | 0.45 | 0.28 | 0.04 |
| **claude3opus** | 0.24 | 0.27 | 0.24 | 0.08 | 0.44 | 0.45 | - | 0.23 | 0.03 |
| **claude3sonnet** | 0.21 | 0.28 | 0.18 | 0.38 | 0.18 | 0.28 | 0.23 | - | 0.19 |
| **claude3haiku** | 0.05 | 0.05 | 0.03 | 0.28 | 0.03 | 0.04 | 0.03 | 0.19 | - |

###### Review 2: Inter-reader variability between the reference and LLMs for review 2

| Interreader variability Kappa of Specific Feature of Interest Detection | | | | | | | | | |
| --- | --- | --- | --- | --- | --- | --- | --- | --- | --- |
|  | gpt_3_5 | gpt_4o | gpt_4turbo | calude3_haiku | calude3_sonnet | calude3_opus | reader_1 | reader_2 | consensus |
| **gpt_3_5** | NA | 0.12 | 0.11 | 0.320 | 0.071 | 0.110 | 0.09 | 0.10 | 0.11 |
| **gpt_4o** | 0.12 | NA | 0.50 | 0.200 | 0.029 | 0.028 | 0.44 | 0.43 | 0.47 |
| **gpt_4turbo** | 0.11 | 0.50 | NA | 0.190 | 0.017 | 0.034 | 0.72 | 0.66 | 0.74 |
| **haiku** | 0.32 | 0.20 | 0.19 | NA | 0.076 | 0.170 | 0.15 | 0.16 | 0.18 |
| **sonnet** | 0.07 | 0.03 | 0.02 | 0.076 | NA | 0.260 | 0.01 | 0.01 | 0.02 |
| **opus** | 0.11 | 0.03 | 0.03 | 0.170 | 0.260 | NA | 0.03 | 0.03 | 0.03 |
| **reader_1** | 0.09 | 0.44 | 0.72 | 0.150 | 0.014 | 0.031 | NA | 0.64 | 0.79 |
| **reader_2** | 0.10 | 0.43 | 0.66 | 0.160 | 0.014 | 0.029 | 0.64 | NA | 0.85 |
| **consensus** | 0.11 | 0.47 | 0.74 | 0.180 | 0.017 | 0.034 | 0.79 | 0.85 | NA |

| Interreader variability Kappa of Abstract Characteristics | | | | | | | | | |
| --- | --- | --- | --- | --- | --- | --- | --- | --- | --- |
|  | gpt_3_5 | gpt_4o | gpt_4turbo | calude3_haiku | calude3_sonnet | calude3_opus | reader_1 | reader_2 | consensus |
| **gpt_3_5** | NA | 0.81 | 0.81 | 0.84 | 0.82 | 0.79 | 0.76 | 0.75 | 0.75 |
| **gpt_4o** | 0.81 | NA | 0.93 | 0.87 | 0.88 | 0.86 | 0.87 | 0.86 | 0.85 |
| **gpt_4turbo** | 0.81 | 0.93 | NA | 0.88 | 0.90 | 0.86 | 0.88 | 0.87 | 0.86 |
| **haiku** | 0.84 | 0.87 | 0.88 | NA | 0.86 | 0.81 | 0.81 | 0.81 | 0.80 |
| **sonnet** | 0.82 | 0.88 | 0.90 | 0.86 | NA | 0.87 | 0.84 | 0.83 | 0.83 |
| **opus** | 0.79 | 0.86 | 0.86 | 0.81 | 0.87 | NA | 0.81 | 0.82 | 0.81 |
| **reader_1** | 0.76 | 0.87 | 0.88 | 0.81 | 0.84 | 0.81 | NA | 0.87 | 0.93 |
| **reader_2** | 0.75 | 0.86 | 0.87 | 0.81 | 0.83 | 0.82 | 0.87 | NA | 0.93 |
| **consensus** | 0.75 | 0.85 | 0.86 | 0.80 | 0.83 | 0.81 | 0.93 | 0.93 | NA |

### S6: LLM price and accuracy

###### Review 1

|  | **GPT-3.5** | **GPT-4T** | **GPT-4o** | **Claude3-Haiku** | **Claude3-Sonnet** | **Claude3-Opus** | **Average across models** | **Total** |
| --- | --- | --- | --- | --- | --- | --- | --- | --- |
| **Prompt 1 input tokens** | 302 | 302 | NA | 340 | 340 | 340 | 324.8 | 1624 |
| **Prompt 2 input tokens** | 148 | 148 | NA | 154 | 154 | 154 | 151.6 | 758 |
| **Prompt 3 input tokens** | 261 | 261 | NA | 273 | 273 | 273 | 268.2 | 1341 |
| **Input tokens per title+abstract*** | 719.7 | 719.7 | 719.7 | 816.0 | 816.0 | 816.0 | 767.8 | 4607 |
| **Output tokens per title+abstract*** | 97.0 | 97.0 | 98.8 | 120.0 | 112.7 | 120.0 | 107.6 | 645,5 |
| **Input prize (USD/million input tokens)** | 1.5 | 10.0 | 5.0 | 0.3 | 3.0 | 15.0 | 5.8 | 34,8 |
| **Output prize (USD/million output tokens)** | 3.0 | 30.0 | 15.0 | 1.3 | 15.0 | 75.0 | 23.2 | 139,3 |
| **Price per abstract (USD/abstract)** | 0.00137 | 0.01011 | 0.00508 | 0.00035 | 0.00414 | 0.02124 | 0.00705 | 0.042 |
| **Prize 500 sample (USD)** | 0.7 | 5.1 | 2.5 | 0.2 | 2.1 | 10.6 | 3.5 | 21,1 |
| **Prize full review (8859) (USD)** | 12.1 | 89.5 | 45.0 | 3.1 | 36.7 | 188.2 | 62.4 | 374,6 |
| *Prompt 1 |  |  |  |  |  |  |  |  |

The figure below illustrates the price in USD and accuracy for the different LLM and human readers.

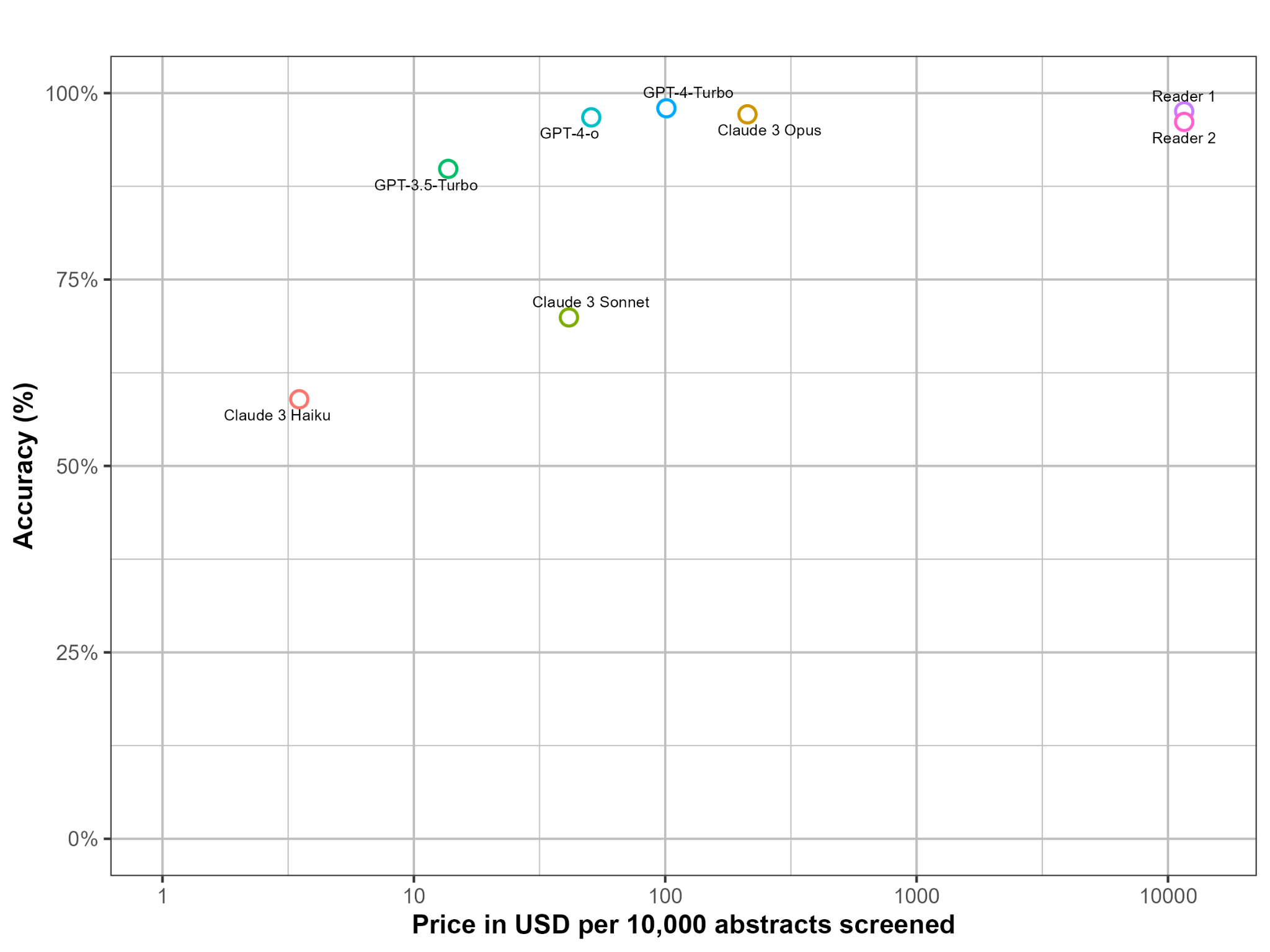

Figure: A semi-log plot of price of screening 10,000 abstracts against accuracy. GPT-4o is the most cost-efficient of the highest-performing LLMs. The human prize is based on 1.7 minutes per title and abstract screening and an hourly salary of 280.2 DKK = 40.94 USD for residents in DK.

### S7: False negative analysis of the 12 included studies in review 1

*Study 1. Development and validation of a deep learning model for brain tumor diagnosis and classification using magnetic resonance imaging. (DOI 10.1001/jamanetworkopen.2022.25608)* was included by both readers and all LLMs, except GPT-3.5, which excluded it from outcome and study type exclusion criteria. However, the abstract explains that it is a diagnostic test accuracy study and that it uses 1 internal and 3 external data sets. It also reports AUC, sensitivity, specificity, and accuracy, so GPT-3.5 seems to have made a mistake in this case.

*Study 2. Deep learning-based computer-aided detection system for automated treatment response assessment of brain metastases on 3d MRI. (DOI 10.3389/fonc.2021.739639)* was included by reader 1, but not reader 2. It was also not included by GPT-3.5 and GPT-4o due to wrong outcome and study type (GPT-3.5) and wrong comparator, outcome, and study type (GPT-4o). In the abstract, it is a diagnostic test accuracy study with an external test set, and it is mentioned that agreements between computer assisted diagnosis (CAD) and radiologists were assessed. For this reason, GPT-3.5 and GPT-4o seem to be wrong here.

*Study 3: Improving the diagnosis of acute ischemic stroke on non-contrast ct using deep learning: a multicenter study. (DOI 10.1186/s13244-022-01331-3)* was included by both readers and all LLMs, except GPT-3.5. It excluded the study due to wrong outcome and study type. However, the abstract mentions AUC and that external validation was performed, and for this reason GPT-3.5 seems to be wrong.

*Study 4: Multidimensional deep learning reduces false-positives in the automated detection of cerebral aneurysms on time-of-flight magnetic resonance angiography: a multi-center study. (DOI 10.3389/fneur.2021.742126)* was included by reader 1, Claude3-Sonnet, and Claude3-Haiku. However it was excluded by reader 2 and the other LLMs due to wrong intervention, comparator, outcome, and study type (GPT-3.5), wrong comparator (GPT-4T), wrong comparator (GPT-4o), wrong comparator, outcome and study type (Claude3-Opus). The abstract explicitly describes the intervention as a CNN, the outcome as sensitivities, and study type as external validation study. However the comparator is not clearly defined, and for this reason, it could be argued that the study should not have been included by reader 1, reference, Claude3-Sonnet, and Haiku.

*Study 5: Diagnostic test accuracy study of a commercially available deep learning algorithm for ischemic lesion detection on brain MRIs in suspected stroke patients from a non-comprehensive stroke center. (DOI: 10.1016/j.ejrad.2023.111126)* was included by both readers and all LLMs.

*Study 6: Brain tumor/mass classification framework using magnetic-resonance-imaging-based isolated and developed transfer deep-learning model. (DOI 10.3390/s22010372)* was included by both readers, GPT-3.5, Claude3-Sonnet and Claude3-Haiku. Meanwhile GPT-4T and GPT-4o excluded it due to wrong comparator, outcome and study type and Claude3-Opus said wrong intervention, comparator, outcome and study type. In the abstract, the intervention is mentioned as a CNN model, the outcome is mentioned as accuracy, and external validation is indirectly written as the model was tested on images from a different machine. However the comparator is not clearly defined, and again it can be argued that the abstract should not have been included by both readers for that reason.

*Study 7: Improvement of automatic glioma brain tumor detection using deep convolutional neural networks. (DOI: 10.1089/cmb.2021.0280)* was included by reader 1, GPT-3.5 and Claude3-Haiku, and excluded by the rest. GPT-4T and GPT-4o excluded due to missing comparator, outcome and study design. The same with Claude3-Opus, which additionally said it should be excluded due to wrong intervention, comparator, outcome and study type. The intervention seems correct in the abstract (CNN for brain tumours), and so does the outcome (accuracy), however comparator and external validation is not clearly stated in the abstract, and for this reason it can be argued that the abstract should have been excluded by the human readers..

*Study 8: Diagnosing autism spectrum disorder in children using conventional mri and apparent diffusion coefficient based deep learning algorithms. (DOI: 10.1007/s00330-021-08239-4)* was included by reader 2 and Claude 3 Haku. All other voted *exclude*, GPT-3.5 due to wrong population, intervention, outcome, and study type. GPT-4T and GPT-4o due to wrong population and missing comparator. Claude3-Opus due to wrong intervention and missing comparator. Claude3-Sonnet due to wrong population and missing comparator. Since the study is on children, it should have been excluded by the human readers due to incorrect population. Moreover, no comparator is clearly defined in the abstract.

*Study 9: Radiological identification of temporal lobe epilepsy using artificial intelligence: a feasibility study. (DOI: 10.1093/braincomms/fcab284)* was excluded by reader 1, GPT-4T (wrong outcome and study type), GPT-4o (wrong population, outcome, and study type) and Claude3-Opus (wrong comparator, outcome and study type). The abstract does explain human interpretation of the scans, but not as the comparator, but as they are scans interpreted as normal by humans. For this reason the comparator is unclear. It does explain the outcome as accuracy, but it is not clear from the abstract that this is an external validation. Due to these inaccuracies, the abstract should probably not have been included.

*Study 10: Pre-trained MRI-based Alzheimer's disease classification models to classify memory clinic patients. (DOI: 10.1016/j.nicl.2020.102303)* was included by reader 1 and the Claude 3 models, while it was excluded by GPT-3.5 (wrong intervention, comparator, outcome and study type), GPT-4T (wrong comparator, outcome) and GPT-4o (wrong comparator). The abstract explains that patients are diagnosed based on their clinical diagnosis and not interpretation of the images, which conflicts with the comparator criteria. The study type does however seem to be an external validation with outcomes of AUC. It can be argued that the study should not have been included due to an inaccurate comparator.

*Study 11: Automatic detection of lesion load change in multiple sclerosis using convolutional neural networks with segmentation confidence. (DOI: 10.1016/j.nicl.2019.102104)* was included by both readers and most LLMs, but excluded by GPT35-T (wrong outcome and study type), GPT-4o (wrong comparator), Claude3-Sonnet (wrong comparator, outcome and study type). The abstract does indicate that it includes an external testset, and that AUC is reported, but the comparator is again not clearly defined. In a more strict interpretation of the criteria it can be argued that this abstract should also not have been included.

*Study 12: Impact of an AI software on the diagnostic performance and reading time for the detection of cerebral aneurysms on time of flight MR-angiography. (DOI: 10.1007/s00234-024-03351-w)* was included by both readers, and all LLMs except GPT-4T (wrong outcome) and Claude3-Opus (wrong intervention). The intervention seems correct in the abstract, as it is an AI-based software trained to detect cerebral aneurysms. Since they tested the software and did not develop it, it seems to be an external test. They also report sensitivity and specificity, and for this reason it seems that the abstract should have been included, and that GPT-4T and Claude3-Opus were wrong here.
